# Aging Out of the Blue: Region-Specific Epigenetic Clock Calibration for a Blue Zone with the DNAm SuperLearner

**DOI:** 10.64898/2026.03.02.26346901

**Authors:** Nolan Gunter, Andres Cardenas, Michael Kobor, Nicole Gladish, Julia L. MacIsaac, Kristy Dever, David Rehkopf, William Dow, Luis Rosero-Bixby, Alan Hubbard

## Abstract

Epigenetic clocks estimate biological age from DNA methylation patterns at CpG sites, providing robust predictions of mortality and morbidity risk. “Blue zones”—regions of exceptional longevity—offer a unique opportunity to investigate how biological aging diverges from chronological age. However, standard clocks are typically trained on large, heterogeneous datasets, reflecting average population trends rather than region-specific dynamics. Using data from the Costa Rican Longevity and Healthy Aging Study (CRELES), we profiled DNA methylation from residents of the Nicoya blue zone (*n* = 206) and a comparison population in other parts of Costa Rica (*n* = 875). We propose training a *SuperLearner*, an ensemble machine learning approach, on the non-Nicoyan Costa Ricans to optimize predictive performance across existing clocks and flexible machine learners. Theoretically justified by its Oracle property, *SuperLearner* performs asymptotically as well as the best candidate predictor in the ensemble, resulting in a weighted combination of algorithms used to predict age. We then used this trained model to construct a calibrated hypothesis test comparing residual age distributions between the blue zone region and the comparison population. Comparing our approach to the five top-performing epigenetic clocks (ranked by MSE) in the Costa Rican cohort, only SuperLearner suggested age deceleration (an average of ∼ 1 year) in the non-Nicoyan reference group. Before calibration, SuperLearner showed the strongest evidence for slowed biological aging among blue zone Nicoyans, estimating a three-year reduction (*R̄*_1_ = −3.05, 95% CI [−3.64, −2.46]) in epigenetic age. Calibrating with non-Nicoyan Costa Ricans improved consistency between estimates in all clocks, decreasing the estimated aging advantage in Nicoyans to about two years (Δ̂ = −1.96, 95% CI [−2.56, −1.37]). This approach provides a robust framework for estimating longevity in distinct regions when a relevant comparison population is available.

## 1 Introduction

The human methylome represents an additional regulatory layer of the genome, in which methyl groups attach to cytosine–guanine dinucleotides (CpG sites) to modulate gene expression, transcription, and other cellular processes. These methylation marks are dynamic, changing systematically with age and environmental exposure. The concept of biological age—a measure intended to capture the physiological state of aging rather than simple time since birth—has emerged as a powerful framework for studying longevity, health, disease and mortality risk. Multiple biomarkers have been proposed to estimate biological age, including those derived from proteomic, transcriptomic, and metabolomic profiles, composite clinical indicators, telomere length, and most prominently, DNA methylation (DNAm)–based “epigenetic clocks” [1]. These clocks use genome-wide methylation data to predict age based on methylation levels at specific CpG sites, producing an estimated DNAm age that reflects the cumulative biological effects of aging processes [2]. DNA methylation–based age estimators have been shown to be among the most accurate and interpretable measures of biological aging [1]. Early approaches, such as Bocklandt’s clock, applied penalized regression models to predict chronological age from methylation levels at selected CpG sites. Among the most promising and widely-used is Horvath’s multi-tissue clock [3], which used elastic-net regression on 353 CpGs in more than 30 tissue and cell types to derive an age predictor, achieving robust performance throughout the human lifespan and with various biological sources. An earlier biomarker, Hannum’s clock [4], trained in adult blood-derived DNA and using 71 CpGs, also demonstrated strong prediction, but showed limited generalizability to other tissues or younger individuals. Subsequent work, including Levine’s “PhenoAge” clock, incorporated clinical biomarkers to capture morbidity and mortality risk in addition to chronological aging [5]. Collectively, these models have established DNA methylation as a consistent and quantifiable correlate of biological aging—one that can be adapted, extended, and tested across many populations.

The term “blue zone”, a region characterized by an exceptionally high number of centenarians, originated in a 2004 study of the island of Sardinia in Italy [6]. Since then, additional blue zones have been identified with researchers eagerly investigating the factors that contribute to such extraordinary longevity [7, 8, 9]. The consensus holds that healthy aging is strongly influenced by the exposome—an ever-evolving network of biological, sociocultural, and environmental factors to which individuals are exposed throughout life [10]. Some epigenetic clocks have been developed in response to these patterns of healthy aging or exceptional longevity. An example is the DNAmFitAge clock, which recognizes the importance of physical activity in modulating biological age and incorporates fitness measures into its predictive model [11]. In contrast, the GrimAge clock was designed to estimate lifespan and healthspan by integrating surrogate DNA methylation markers for plasma proteins and smoking history, making it one of the most powerful predictors of mortality risk to date [12]. More recently, Centenarian clocks have been developed to validate claims of exceptional longevity by identifying unique methylation signatures for individuals who have surpassed the centenarian threshold [13].

Standard practice in investigating patterns in epigenetic-aging research typically proceeds in two steps: first, a fixed clock model is used to predict chronological age from DNA methylation data; second, an age-acceleration metric is derived, usually by taking either the raw difference between DNA methylation–based age and chronological age or the residual from regressing epigenetic age on chronological age. These residuals quantify age deceleration or perhaps acceleration, denoting the amount to which an individual is biologically younger (or older) than expected for their chronological age. Longevity research extends this reasoning to extreme populations—centenarians of high-longevity regions frequently show lower mean epigenetic age than expected, and these age decelerations are widely taken as evidence of exceptional biological aging.

However, although fixed-clock residuals are widely used, they may not be optimal for detecting the effects of biological aging in such exceptional regions. Most epigenetic clocks are trained on large, heterogeneous populations, thus capturing average methylation–age relationships across many cohorts [14]. For instance, the Horvath clock is trained on almost 40 data sets, then tested on over 30 data sets [3]. This pooled training encourages models to capture global average methylation–age relationships rather than region–specific dynamics that might be influenced by local lifestyle and exposures. In our motivating example, we address this by utilizing a dedicated comparison population to calibrate the epigenetic model. This allows inferences to be drawn from a localized comparison between a “blue zone” and a control group that is not reported to have unusual longevity. While the choice of this reference group is critical to the ultimate interpretation, leveraging a population that shares certain baseline traits with the blue zone enables a more refined analysis of epigenetic aging. We acknowledge, however, that if the comparison population possesses unrecognized longevity-promoting characteristics similar to the blue zone, this proximity may lead to more conservative estimates, potentially biasing results toward the null.

To detect biological deviations in atypical regions, the use of an appropriate reference group offers a complementary and potentially more robust approach to traditional aging metrics. Rather than relying on fixed global clocks, we propose training and calibrating an epigenetic model on a well-matched non-blue zone reference group, then applying the calibrated predictor to the blue zone cohort. Training using a population with similarities to the blue zone removes spurious calibration differences between-population and yields a test sensitive only to aging residual differences that persist after calibration. In doing so, we can more confidently attribute any observed age-deceleration signal to authentic region–specific longevity effects—rather than artifacts of model training or population mismatch. Ultimately, our approach provides a principled basis for evaluating claims of exceptional aging in blue zones.

Section II introduces the SuperLearner framework used to construct epigenetic age predictors. We detail how region-specific calibration corrects systematic misalignment between methylation and chronological age and improves power for detecting true biological deviations across populations. Section III presents an empirical case study using data from a nationally representative longitudinal study of the Costa Rican population ^1^, which included oversampling of the older population located on the Nicoya peninsula—one of the best documented blue zones in the world [15]. We apply the calibrated SuperLearner approach to compare biological aging between Nicoyan and non-Nicoyan Costa Ricans, illustrating how calibration alters inference in practice. Section IV concludes with implications for blue zone and longevity research. We discuss how calibration reframes cross-population comparisons, and provides a principled pathway for evaluating biological age across genetically and culturally distinct populations when testing the impact of an exposure.

## 2 Methods

### 2.1 DNA Methylation

Whole blood was collected and processed as previously described [16]. 750 ng of extracted genomic DNA was bisulfite converted according to manufacturer’s instructions (Zymo EZ DNA Methylation Kit, Zymo Research, Irvine, CA). Samples were then randomized in a stratified manner to ensure even distribution across plate, row, and chip and 160 ng of converted DNA was amplified and hybridized to the Illumina MethylationEPICv1 BeadChip arrays (Illumina Inc., San Diego, CA) following standard protocols.

To improve computational efficiency and ensure biological comparability across array platforms, we restricted analyses to CpG sites consistently measured across all generations of Illumina methylation arrays. While the full Illumina EPIC v2 array measures over 850,000 CpGs, earlier arrays such as the 450K and EPIC v1 include smaller and partially overlapping subsets. Many CpGs unique to a given platform are either poorly replicated, platform-specific, or absent in archival datasets, limiting interpretability and reproducibility across studies. Then, to retain only universally profiled CpGs, we identified the intersection of loci annotated in the Illumina 450K, EPIC v1, and EPIC v2 annotation packages available through *Bioconductor* in R. Using these annotations, we obtained the common probe identifiers and restricted the methylation matrix to the resulting overlapping set of 350,476 CpG sites. This filtering step eliminates roughly half of the probes from the full array while preserving biologically informative regions represented across all array generations. Reducing the analysis to these shared CpGs not only mitigates memory constraints but also enhances generalizability of downstream models by anchoring results to loci reproducibly captured in historical and contemporary methylation assays.

We further reduced the dimensionality of the methylation matrix by applying a variance-based feature selection step. CpG sites with negligible variability across individuals contribute little to model fitting or biological signal detection, while substantially increasing computational cost. To remove such uninformative loci, we computed the empirical variance of methylation beta values for each CpG site across all samples and excluded those with variance below 0.005. This conservative threshold retains probes exhibiting meaningful inter-individual methylation differences while discarding sites that are constitutively methylated or unmethylated in nearly all samples. After this filtering, 89,407 CpG sites remained, representing the most variable and potentially biologically informative subset of the methylome for subsequent modeling. We also retain any CpG used for the epigenetic clocks that we implement in a separate CpG set. For instance, the Horvath clock uses 353 fixed CpG sites, and we retained as many of these as were available in the original data.

### 2.2 Ensemble Epigenetic Clocks through SuperLearner

*SuperLearner* is an ensemble machine learning algorithm that provides a theoretically optimal method to combine the predictions of multiple models into a single data-adaptive estimator [17]. Rather than assuming that any one model captures the true underlying relationship between predictors and outcome, SuperLearner seeks to approximate the best possible predictive function from a convex combination of candidate learners. This framework is particularly useful for modeling complex biological processes, such as DNA methyla-tion–age relationships, where the signal arises from thousands of CpG sites and no single parametric model is likely to be correct. SuperLearner has been used to optimize epigenetic age before, such as in one study that constructed a sperm clock for men to predict probability of pregnancy in couples with penalized regressions like LASSO, ridge, elastic net and also multivariate adaptive regression splines [18]. The SuperLearner framework was recently expanded into a standardized pipeline that incorporates principal component analysis (PCA) to manage high-dimensional CpG data, demonstrating that ensemble-based predictors significantly improve accuracy over traditional single-model approaches [19].

Users must first add each algorithm to a pre-defined “library”—for example, a linear model, random forest, elastic net, gradient boosting machine, or a fixed epigenetic clock such as Horvath’s or GrimAge. Each of these is then fit using *K*-fold cross-validation to produce out-of-sample predictions. These cross-validated predictions are combined through a meta-learner, typically a convex combination estimated by least squares regression. The resulting weighted combination of algorithms is the *ensemble* Super Learner, while the single best-performer (based on cross-validated risk) is termed the *discrete* Super Learner. This structure allows the method to balance bias and variance across diverse learners, weighting algorithms higher when they optimize the specified loss function and improve predictive accuracy.

Theoretical guarantees make SuperLearner distinct from *ad hoc* ensembling. Under mild regularity conditions, van der Laan, Polley, and Hubbard [17] proved that the cross-validated Super Learner achieves the same asymptotic performance as the best possible convex combination of the candidate algorithms—a result known as the Oracle inequality. In practice, this means that as sample size grows, SuperLearner performs as well as if one had known in advance which single model or combination of models would minimize true prediction error. This property holds irrespective of whether the best learner is parametric, semi-parametric, or nonparametric, making the approach well suited for biological data with complex nonlinear relationships. While SuperLearner has been used in epigenetic aging prediction before, it has never been used in a way that combines well-established epigenetic clocks with adaptable machine learning algorithms. Fixed clocks such as Horvath’s multi-tissue model [3], Hannum’s blood-based clock, Levine’s PhenoAge [5], or Lu’s GrimAge [12] can each be treated as distinct learners within the SuperLearner library. Alongside these, flexible algorithms like random forests, support vector machines, and gradient boosting, can capture residual patterns related to age in the comparison group not fully explained by fixed clocks. The ensemble then provides both (i) an oracle-weighted combination representing the best possible integrated predictor, and (ii) identification of the single best performing learner within the library (discrete SuperLearner). This makes the SuperLearner framework a strong choice for studying region-specific dynamics of biological aging, where the optimal combination of clocks and machine learning models may differ from the more diverse population in which the fixed clocks were trained. For more discussion on SuperLearner, we refer to [20], and in particular emphasize the second figure of the paper for a detailed schematic description of the algorithm. Figure 1 below provides a visual summary of how the training of the SuperLearner works in the context of epigenetic age prediction.

**Figure 1:**
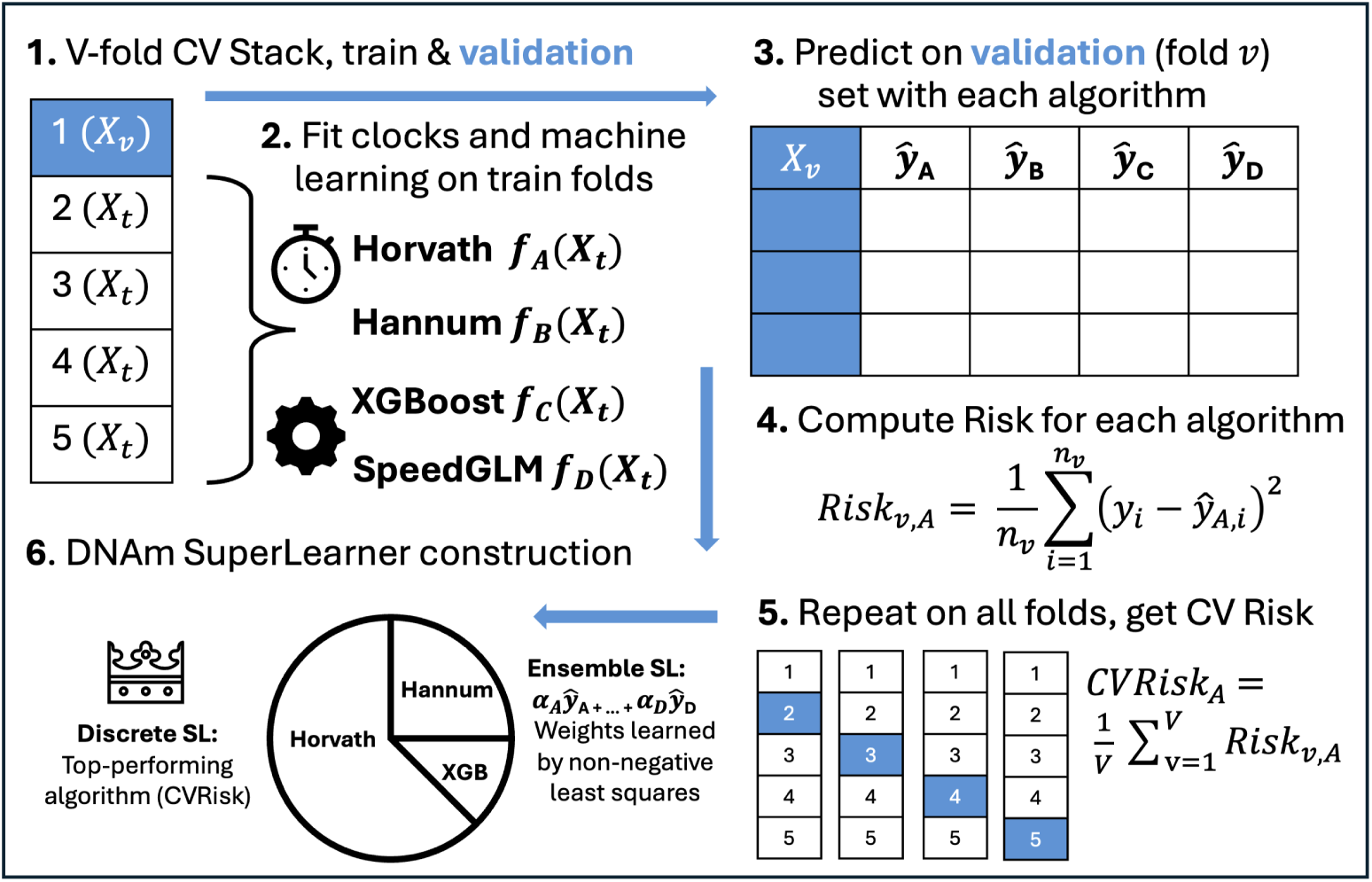
Visual step-by-step summary of the DNAm SuperLearner algorithm. First, the data is split into V-folds. The epigenetic clocks and other machine learning algorithms are fit on the training folds, then used to predict with data from the validation (left out) fold. The risk function (MSE in the figure) is then calculated for each algorithm in the validation set, using the true outcomes. This is iterated V-times, and then the cross-validated risk (CVRisk) is calculated for each algorithm. The discrete SuperLearner is the algorithm with the best CVRisk, and the ensemble is a meta-learner of the algorithms with weights learned by non-negative least squares. In the example, we presents the algorithm library with two clocks (Horvath’s and Hannum’s) and two machine learners (XGBoost and SpeedGLM).

### 2.3 Calibration and Statistical Power

Let *A* ∈ {0, 1} denote membership in the reference (comparison/control) population (*A* = 0) versus the blue zone (or exposed/treatment) population (*A* = 1). For subject *j* in population *a* 0, 1 , let *Y*_aj_ represent chronological age, *W*_aj_ ∈ ℝ^p^ denote methylation features (e.g., CpG levels), and *f* (.) be an epigenetic clock that predicts age from *W* . A fixed “global” clock (such as the Horvath clock) is typically trained on pooled heterogeneous data. In contrast, we define a reference-calibrated clock as the function

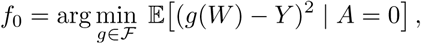

which minimizes prediction based on the mean squared error (MSE) for the reference population only. The resulting minimizer, *f̂*_0_(*W* ) = 𝔼̂ (*Y* |*A* = 0*, W* ) represents the optimal predictor of chronological age within the comparison cohort. In practice, *f*_0_ is estimated using a SuperLearner ensemble. To ensure the clock is unbiased for the comparison group, we perform a simple mean calibration. Let residuals in the comparison/reference (*A* = 0) and blue zone (*A* = 1) data be defined respectively as:

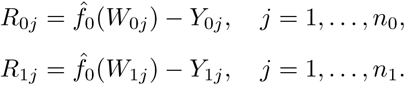

Note that the residuals of individuals within the blue zone are calculated from a prediction function trained *only* on the comparison, *f̂*_0_. We compute the average residual among the reference population and adjust the model predictions by this constant shift:

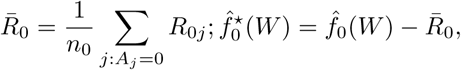

where *f̄*^*^_0_(*W* ) is the re-calibrated learner for the comparison population, so that the mean residual among comparison individuals satisfies

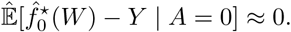

The estimand of interest is the population shift in mean residuals, which under proper calibration (E[*R*_0_] = 0) simplifies to a single term,

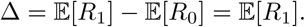

We estimate Δ empirically with Δ̂ and test for a systematic deviation in biological aging using a one-sample *t*-test:

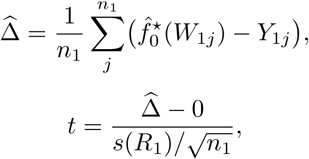

where *s*(*R*_1_) is the sample standard deviation of the residuals of the blue zone. The null hypothesis *H*_0_ : Δ = 0 corresponds to no population-level deviation in biological aging, while the alternative *H*_1_ : E[*R*_1_] *<* 0 (or *>* 0) tests for systematically lower (or higher) predicted biologic ages relative to chronological age. A standard way to evaluate the difference might be to perform a two-sample t-test on the residuals of each population from a fixed epigenetic clock. However, this can inflate bias because the clock’s baseline error is not shared equally across groups—its intercept and scaling are learned from the comparison population only, meaning the residual distribution in the target population reflects both biological differences and systematic miscalibration. This issue arises because it is reasonable to assume that each population has its own optimal epigenetic regression function. A single global clock therefore contains within-population error (noise in predicting individuals) and between-population error (systematic misfit due to differences in age–methylation structure across groups). By training the clock exclusively on a large matched comparison group, we eliminate the between-population component by construction and interpret deviations in the target population solely as departures from the learned reference aging process.

The mean-centered calibration test thus provides a principled way of disentangling genuine regional (or exposure) differences from model misspecification. When a consistent negative (or positive) residual shift is observed after local reference calibration, it more robustly reflects a population-level deviation in aging rate rather than mismatch in the global regression function. A short heuristic example (Appendix) illustrates this decomposition in a toy setting, showing how fitting a reference-based clock removes between-population error while preserving biologically meaningful deviations.

### 2.4 SuperLearner Architecture

The SuperLearner ensemble was constructed from a diverse library of candidate learners spanning both fixed epigenetic clocks and machine learning algorithms with different predictor-screening strategies. This design allows the ensemble to integrate biologically grounded predictors with data-adaptive models capable of capturing residual methylation–age signal.

- **Fixed epigenetic clocks:** Horvath [3], Hannum [4], Levine (PhenoAge) [5], Horvath’s Skin and Blood [21], Elastic Net [22], ENCen40 [13], DNAmGrimAge2 [23], PCGrimAge [24], and DunedinPACE [25] clocks were included as standalone learners. Each clock provides a biologically interpretable predictor of epigenetic age derived from established CpG subsets and regression coefficients published in prior studies. Including these fixed clocks in the library ensures that the ensemble can recover or improve upon existing benchmark models of aging. We used the implementation as part of the methylclock package in Bioconductor and the dnaMethyAge [26] for the first five clocks and for PCGrimAge. Then, for ENCen40, GrimAgeV2, and DunedinPACE, we replicated the algorithms as new learners using all available CpG sites. Where CpG sites were not available in the data, we imputed a 50% methylation value across all observations and used it with the coefficient in the fixed learner.
- **Machine learning algorithms:** We chose data-adaptive learners that are well-suited for handling high-dimensional covariates, including elastic net regression (glmnet), ridge regression, random forest (ranger), gradient boosting (xgboost), fast linear regression (speedglm), and high-dimensional penal-ized regression (biglasso). These models were trained on the full filtered CpG matrix or on feature subsets defined by screening procedures described below.
- **Predictor-screening strategies:** To manage very high-dimensional input and improve computational tractability, each algorithm was paired with a variable-screening methods. The primary screeners included:
- **–** `screen.corP.min25`: selects CpG sites based on the p-values from marginal Pearson correlation tests against chronological age. We retained all sites meeting a pre-specified significance threshold ((*p* ≤ 0.1)); however, to ensure stable model performance in lower-signal environments, the algorithm was constrained to retain at least the top 25 most significantly correlated sites, regardless of the p-value threshold.
- **–** `screen.glmnet.min25:` selects CpG sites with non-zero coefficients from an elastic net model using the *λ* that minimizes mean cross-validated error, *λ*_min_. If *λ*_min_ yields fewer than 25 features, the regularization penalty is relaxed along the solution path to the first *λ* that retains at least 25 CpG sites.
- **–** `screen.rf.clock:` selects CpG sites based on a Random Forest model with 300 trees. Features are selected based on Gini importance (node impurity), retaining the top *n* = 1000 sites.
- **–** screen.clock_all: uses all CpGs included in the fixed clocks combined, except for PCGrimAge, as it uses over 80,000 CpGs.

Each learner produced out-of-sample predictions through *K* = 5-fold cross-validation. These were combined in a convex meta-learner estimated via least squares regression, allowing the ensemble to adaptively weight both fixed and data-driven components based on predictive performance.

## 3 Results

The Costa Rican Longevity and Healthy Aging Study (CRELES) follows a nationally representative cohort of older adults in Costa Rica. The baseline sample includes 2,827 individuals aged 60 and above who were initially surveyed between 2004 and 2006, with a follow-up wave conducted during 2006–2008. A random sample of *n* = 1081 participants from the CRELES cohort were selected for blood sampling for DNA methylation (DNAm) analysis, with about half being sampled in 2017 and half in 2024. In addition, CRELES oversampled data from a complementary group of exceptionally long-lived individuals from the Nicoya region, specifically residents aged 95 years or older. Therefore, we restrict the interpretation of our epigenetic aging estimates to Costa Ricans above the age of 60. Further details about the CRELES study have been described here [27, 28, 29].

### 3.1 Descriptive Analyses

We trained the SuperLearner model on only the reference non-Nicoyan group with *n* = 875 individuals, resulting in the following ensemble SuperLearner in Table 1. Four fixed epigenetic clocks were chosen, with the PCGrimAge clock getting 73.6% of the weight and the Hannum clock 16.1%. Fixed elastic net clocks were also given weight, and a very small portion (0.7%) went to ridge regression that used all CpG sites from the fixed clocks. The discrete SuperLearner is the PCGrimAge algorithm, but for our analyses we use the ensemble.

**Table 1:** Super Learner ensemble weights (nonzero), trained on the Non-Nicoyan reference/comparison group from Costa Rica (*n* = 875), with four established epigenetic clocks and ridge regression chosen.

| Learner | Weight |
| --- | --- |
| SL.PCGrimAge | 73.6% |
| SL.Hannum | 16.1% |
| SL.EN | 7.9% |
| SL.ENCen40 | 1.7% |
| SL.ridge.screen.clock.all | 0.7% |

To compare the results of the calibrated SuperLearner, we compute the same calibrations for five fixed epigenetic clocks. We had access to a comprehensive dataset with most well-known epigenetic clocks, and chose the five with the lowest mean squared error (MSE) across both non-Nicoyans and Nicoyans. This resulted in DNAmFitAge, DNAmGrimAge2, PCGrimAge, ENCen40, and NNCen40. The latter two centenarian clocks have actually been partially trained using about 45% of the CRELES data. Thus, they have an advantage compared to other fixed clocks (and SuperLearner) but may experience some overfitting bias. Figure 2 shows the MSE for each clock, decomposed into the bias squared and variance. The SuperLearner achieves the lowest MSE (16.1), with the next best performing clock being the NNCen40 (MSE = 25.2). Next, we plot the residual distributions in the reference/comparison population and Nicoya in Figure 3. SuperLearner is the only clock that suggests age deceleration (*R̄*_0_ = 1.09) in the reference population.

**Figure 2:**
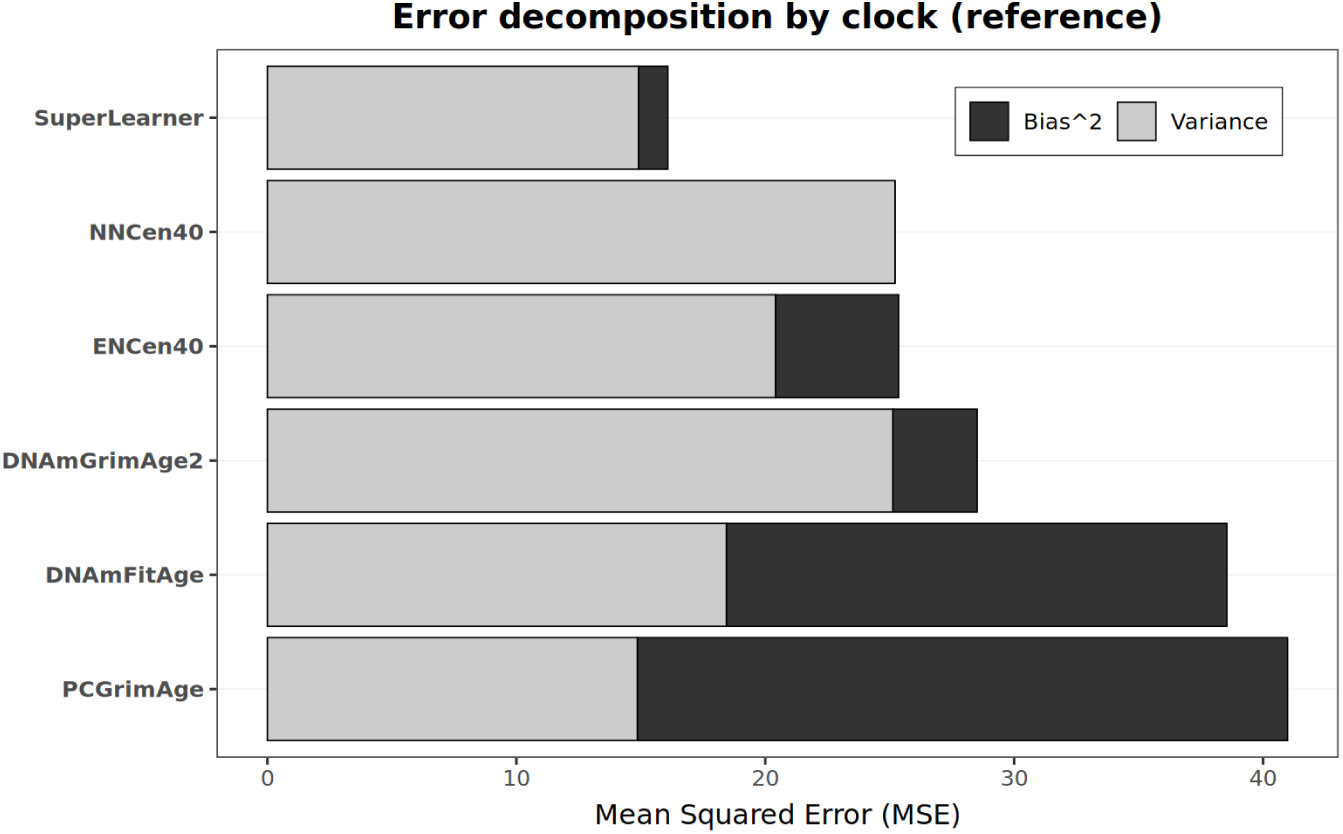
Mean Squared Error (MSE) decomposition for the SuperLearner and the five fixed epigenetic clocks chosen for comparison based on performance in the reference population outside of Nicoya. Grey (light) bars indicate the total variance for the clock, and black (dark) bars indicate the squared. The sum of these two creates the total MSE, which is shown on the horiztonal axis.

**Figure 3:**
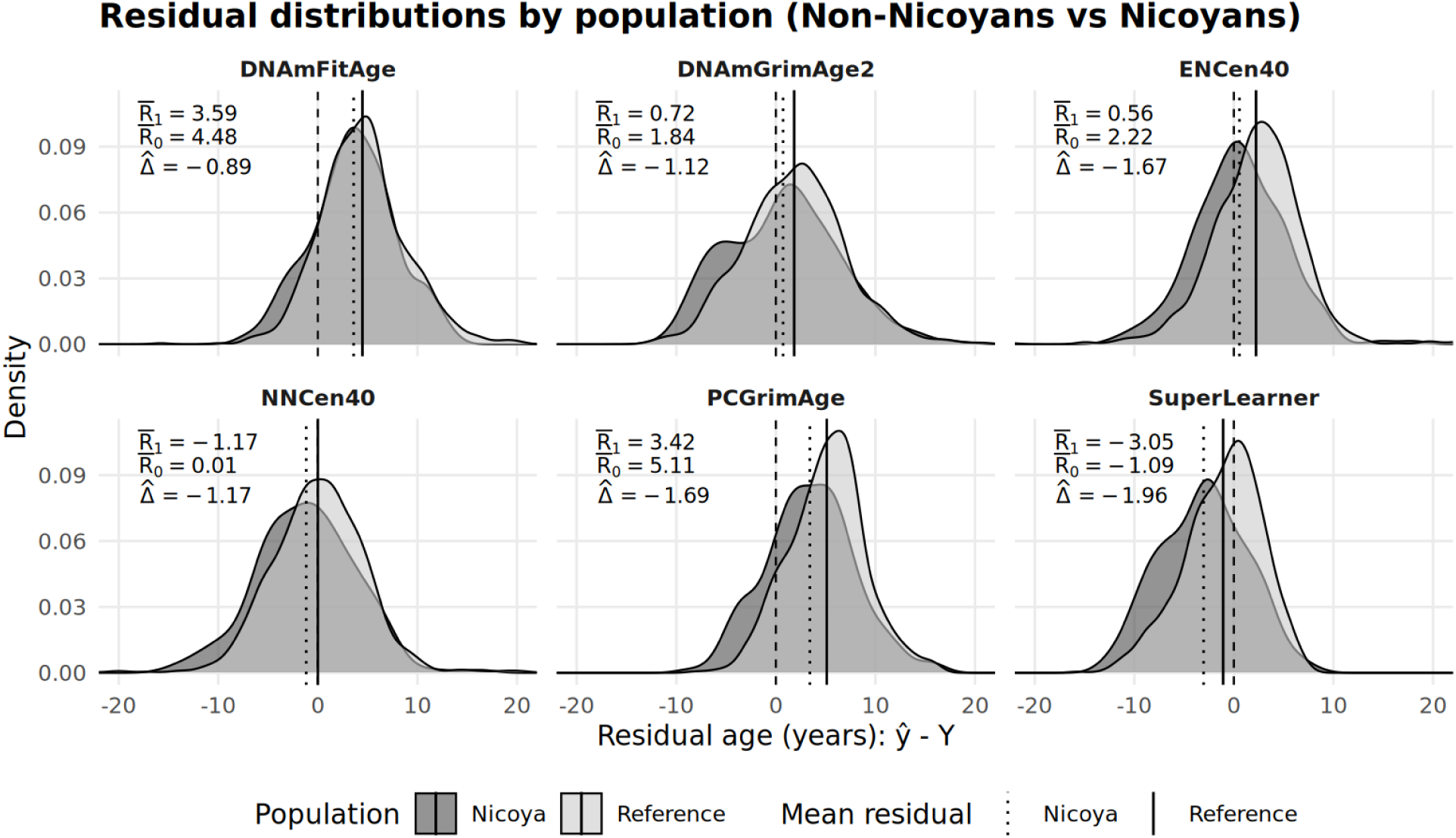
Residual age distributions (*ŷ Y*) for the non–Nicoya reference population (light grey) and Nicoya (dark grey) across five fixed epigenetic clocks and the Super Learner. Each panel shows a histogram of residuals with a vertical line marking the mean residuals in the reference *R̄*_0_, and a vertical dotted line showing the residual mean in Nicoya *R̄*_1_. Δ̂ represents the difference in these two values, or the calibrated estimate of epigenetic aging in Nicoya, indicated by the dashed vertical line. Negative values indicates underprediction of chronological age (younger epigenetic age), while positive indicate accelerated aging.

Instead, most of the other clocks suggest an age acceleration in the reference population (positive residuals on average). The difference in distributions can give a rough visual of the predicted aging differences between populations. Finally, we plot true age vs predicted age in each clock for Non-Nicoyans and Nicoyans in Figure 4.

**Figure 4:**
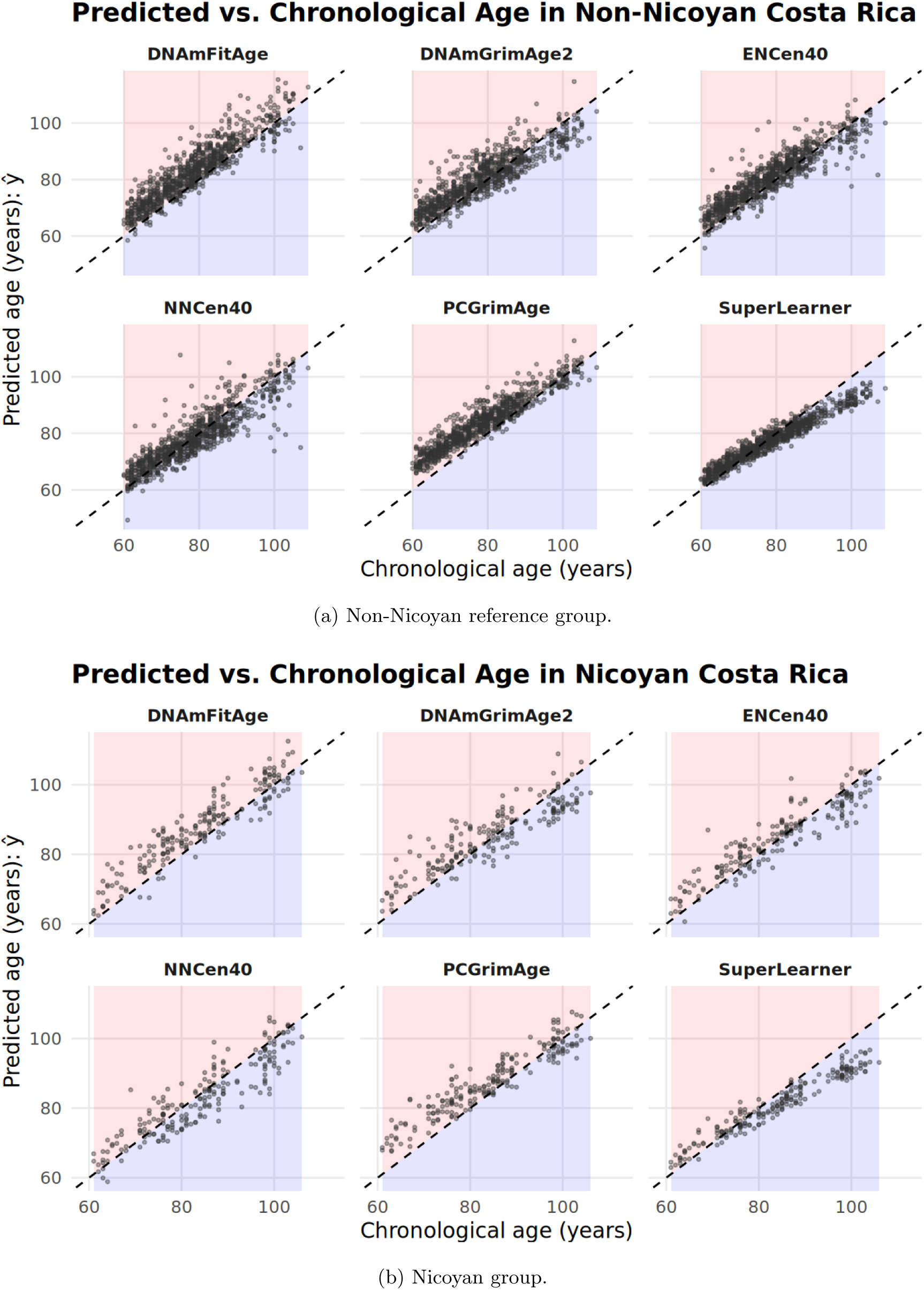
Chronological vs. predicted age in the non-Nicoyan reference group and in Nicoya. Predicted age is unadjusted and uncalibrated on the y-axis for five fixed epigenetic clocks and the Super Learner. The dashed diagonal line at marks predicted age matching chronological age. If a point falls below this (blue shading), it suggests healthy or slowed biological aging. If above (red shading), then it suggests accelerated aging. These panels visualize age-dependent structure prior to any adjustments, showing the baseline for how model calibration occured for the Nicoyan population.

### 3.2 Calibration Results

In Figure 5 we display the 95% confidence intervals of the mean residual epigenetic age for the five top performing fixed clocks and the SuperLearner trained on non-Nicoyan Costa Ricans. For each clock, we plot five confidence intervals using various estimation methods which we describe below. As a baseline, we compute a one-sample t-test on the uncalibrated residuals for each clock (Table 2), which we denote as a naive estimate. Note that we refer to *R̄*_1_ as the uncalibrated mean residual, and Δ̂ as the calibrated. Before calibration, the SuperLearner shows the most evidence for potential age deceleration among Nicoyans aged 60 and older, with a mean residual of *R̄*_1_ = 3.05, 95% CI [ 3.64, 2.46]. This translates to an estimated average of three epigenetic years less than chronological age in older Nicoyans. The NNCen40 clock also suggests this, but with a near three-times lower aging advantage *R̄*_1_ = 1.17, 95% CI [ 1.85, 0.49]. The other uncalibrated clocks suggest much different results, with ENCen40 and DNAmGrimage2 showing no significant difference, and PCGrimAge and DNAmFitAge actually estimating mean age deceleration in Nicoya. Clearly, the choice of epigenetic clock greatly impacts the results of such a hypothesis test with uncalibrated residuals.

**Figure 5:**
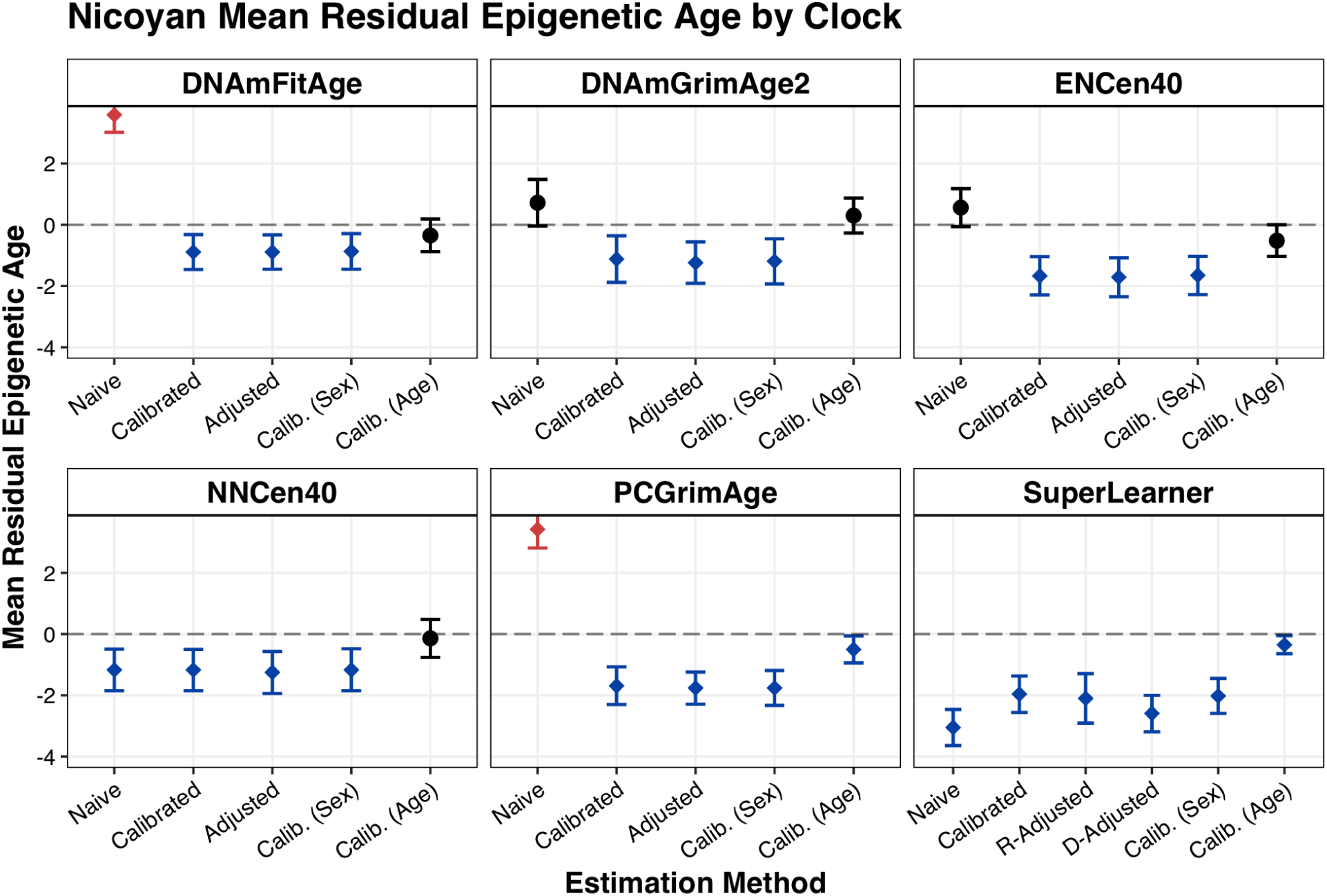
95% confidence intervals for mean residual age estimates in Nicoya, stratified by clocks and associated estimation methods. Confidence intervals below the dashed zero-line (blue) suggest statistically significant evidence for decelerated biological aging (negative residual difference between predicted age and chronological age). Confidence intervals above (red) suggest accelerated biological aging. Naive estimation denotes the uncalibrated, unadjusted residuals (see Table 2. Calibrated denotes subtracting the mean predicted epigenetic age residual in the comparison (non-Nicoyans) from the Nicoyan residuals (see Table 3)). Adjusted denotes running the results with a regression against a confounder set, with R- and D-Adjusted denoting necessary modifications for the SuperLearner (see Appendix). Further calibrations were denoting using biological sex and 10-year binned age subgroups, see Appendix.

**Table 2:**
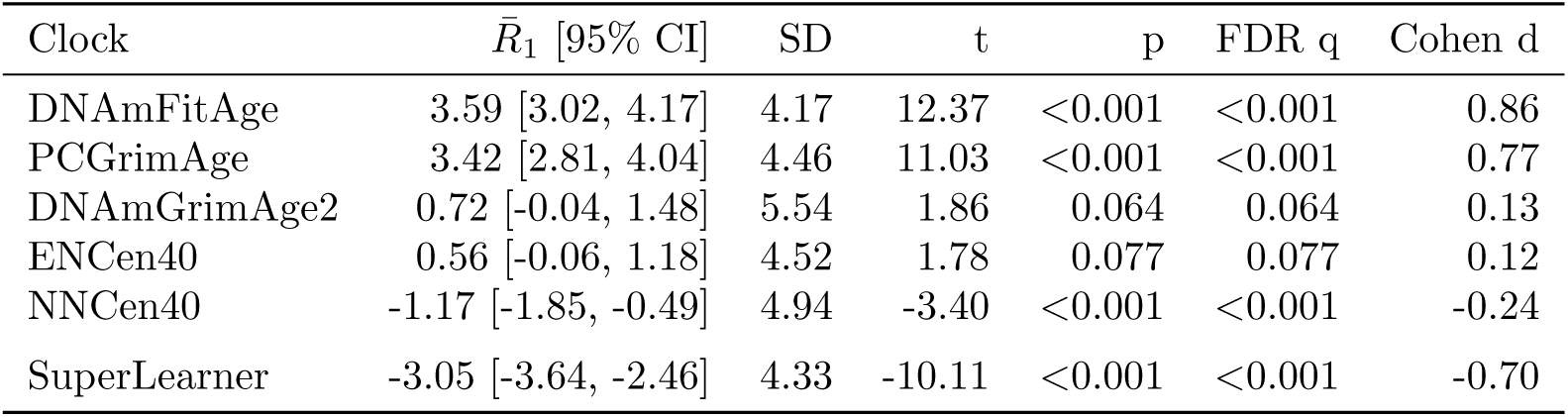
One-sample t-tests in Nicoya of *naive* (uncalibrated) residuals (*H*_0_ : *R̄*_1_ = 0)

| Clock | $\bar{R}_1$ [95% CI] | SD | t | p | FDR q | Cohen d |
| --- | --- | --- | --- | --- | --- | --- |
| DNA<sub>m</sub>FitAge | 3.59 [3.02, 4.17] | 4.17 | 12.37 | <0.001 | <0.001 | 0.86 |
| PCGrimAge | 3.42 [2.81, 4.04] | 4.46 | 11.03 | <0.001 | <0.001 | 0.77 |
| DNA<sub>m</sub>GrimAge2 | 0.72 [-0.04, 1.48] | 5.54 | 1.86 | 0.064 | 0.064 | 0.13 |
| ENCen40 | 0.56 [-0.06, 1.18] | 4.52 | 1.78 | 0.077 | 0.077 | 0.12 |
| NNCen40 | -1.17 [-1.85, -0.49] | 4.94 | -3.40 | <0.001 | <0.001 | -0.24 |
| SuperLearner | -3.05 [-3.64, -2.46] | 4.33 | -10.11 | <0.001 | <0.001 | -0.70 |

**Table 3:** One-sample t-tests in Nicoya of *calibrated* residuals (*H*_0_ : Δ̂ = 0)

| Clock | $\hat{\Delta}$ [95% CI] | SD | t | p | FDR q | Cohen d |
| --- | --- | --- | --- | --- | --- | --- |
| DNA<sub>m</sub>FitAge | -0.89 [-1.46, -0.32] | 4.17 | -3.06 | 0.003 | 0.003 | -0.21 |
| DNA<sub>m</sub>GrimAge2 | -1.12 [-1.88, -0.36] | 5.54 | -2.91 | 0.004 | 0.004 | -0.20 |
| NNCen40 | -1.17 [-1.85, -0.50] | 4.94 | -3.41 | <0.001 | <0.001 | -0.24 |
| ENCen40 | -1.67 [-2.29, -1.04] | 4.52 | -5.28 | <0.001 | <0.001 | -0.37 |
| PCGrimAge | -1.69 [-2.30, -1.07] | 4.46 | -5.43 | <0.001 | <0.001 | -0.38 |
| SuperLearner | -1.96 [-2.56, -1.37] | 4.33 | -6.51 | <0.001 | <0.001 | -0.45 |

**Table 4:**
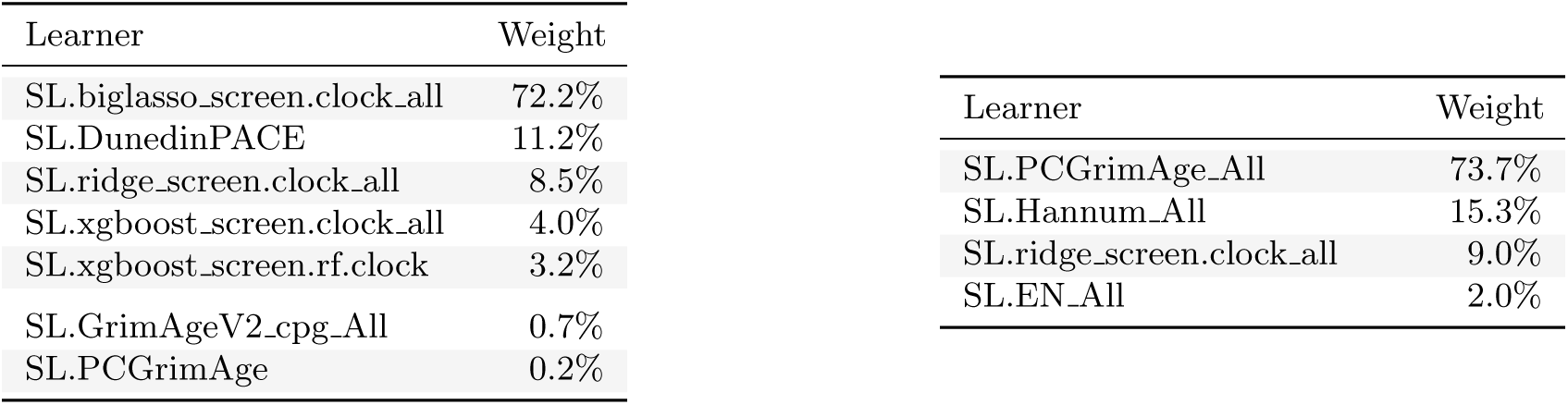
Ensemble Weights for the Residual-Adjusted SL (Left) and Direct-Adjusted SL (Right)

| Learner | Weight |  | Learner | Weight |
| --- | --- | --- | --- | --- |
| SL.biglasso.screen.clock_all | 72.2% |  | SL.PCGrimAge_All | 73.7% |
| SL.DunedinPACE | 11.2% |  | SL.Hannum_All | 15.3% |
| SL.ridge.screen.clock_all | 8.5% |  | SL.ridge.screen.clock_all | 9.0% |
| SL.xgboost.screen.clock_all | 4.0% |  | SL.EN_All | 2.0% |
| SL.xgboost.screen.rf.clock | 3.2% |  |  |  |
| SL.GrimAgeV2.cpg_All | 0.7% |  |  |  |
| SL.PCGrimAge | 0.2% |  |  |  |

Finally, we conduct the same one-sample t-tests but using the calibrated results, that is; subtracting the mean residual for each clock from the reference population from the residuals in Nicoya (Table 3. After calibration, the results become much more consistent across the clocks, all suggesting some level of age deceleration in older Nicoyans. The SuperLearner still estimates the largest age deceleration with a calibrated mean residual of Δ̂ = −1.96, 95% CI [−2.56, −1.37]. The PCGrimAge has the next highest estimate (Δ = 1.69, 95% CI [ 2.30, 1.07]), which aligns with SuperLearner choosing three-quarters of its weight with this clock.

In the Appendix, we also include confounder-adjusted, rather than calibrated, results (see Table 5). We regress the predictions of each clock on four confounders that may contribute to epigenetic aging: sex, body mass index, smoking and socioeconomic status. The results for each clock are similar to that of calibration, with the main SuperLearner estimate being (Δ = 2.10, 95% CI [ 2.91, 1.29]). Additionally, we explore a more fine-tuned calibration strategy using subgroups. Instead of calibrating with the overall sample mean, we calibrate to the means of important subgroups (men/women and 10-year age groups). We find that calibrating to biological sex has little effect on the estimates (Table 6), while calibrating to the 10-year age bins removes most of the effect estimates (Table 7).

**Table 5:**
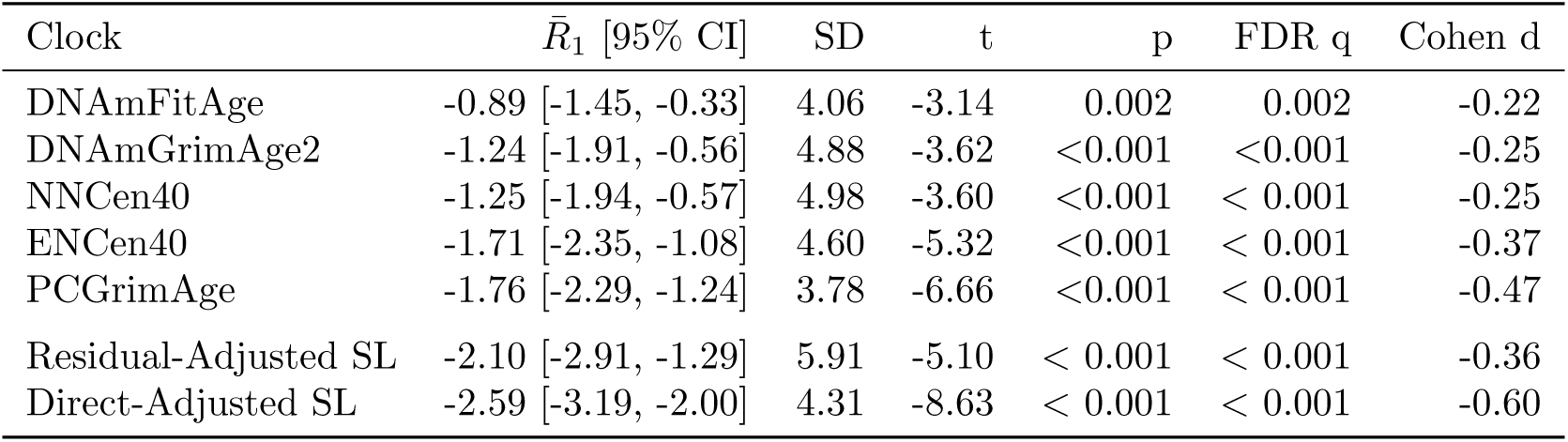
Nicoya: One-sample t-tests of *covariate-adjusted* residuals (Controlled for Sex, BMI, SES proxy and Smoking)

**Table 6:**
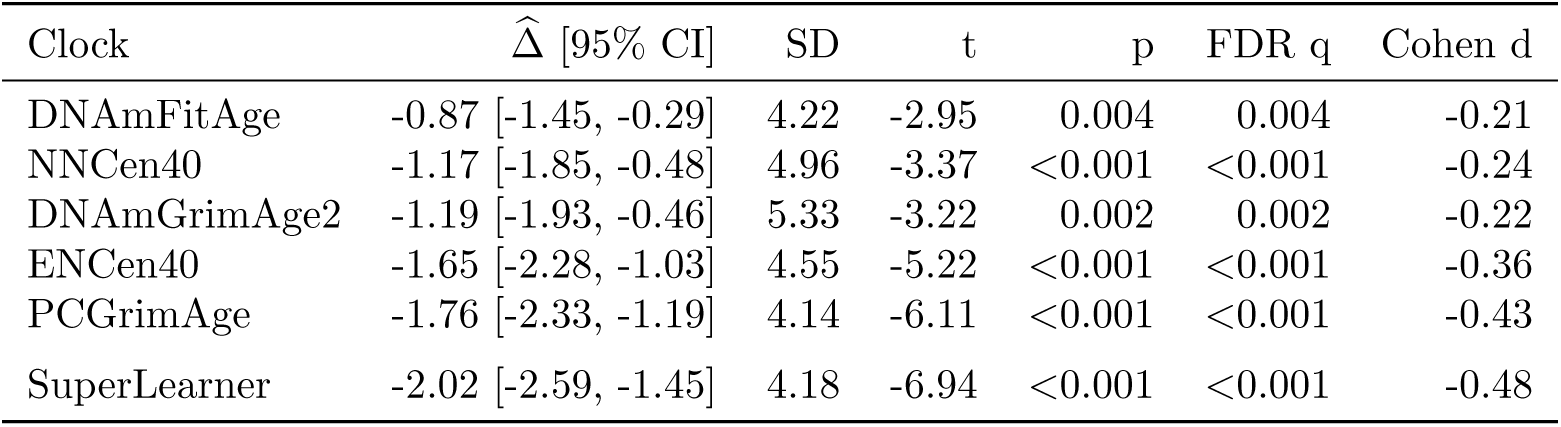
One-sample t-tests in Nicoya of *sex subgroup-calibrated (TMLE)* residuals (*H*_0_ : Δ̂ = 0)

**Table 7:**
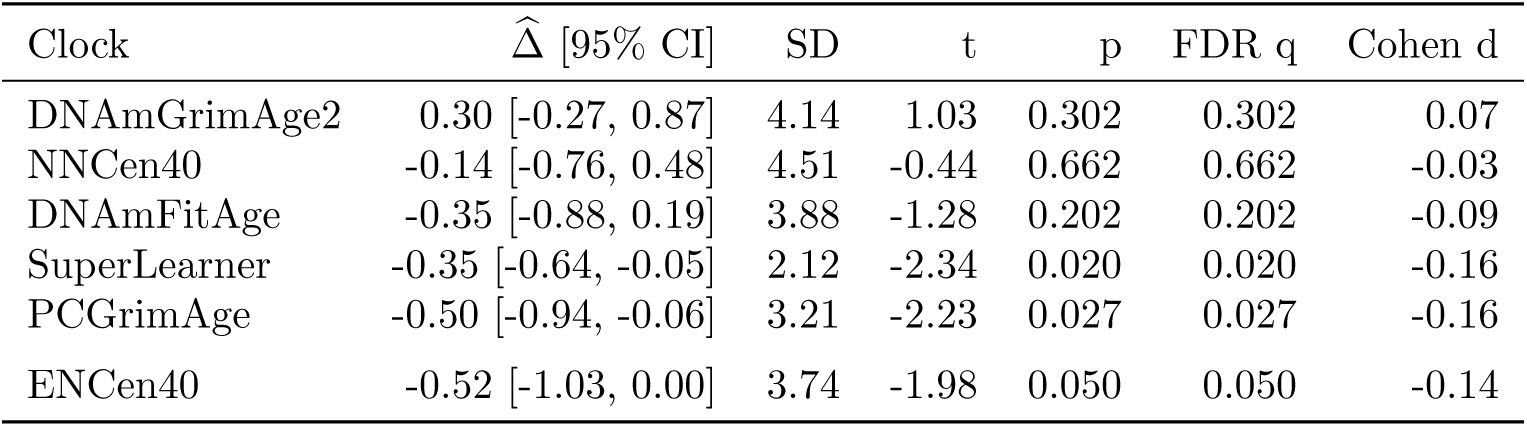
One-sample t-tests in Nicoya of *subgroup-calibrated (TMLE)* residuals with 10-year age-bins from 60 to 110 (*H*_0_ : Δ̂ = 0)

## 4 Discussion

We introduced a novel, robust framework to predict epigenetic aging using SuperLearner, combining well-established fixed epigenetic clocks and flexible machine learning algorithms. This meta-learning approach can be used for optimal learning of epigenetic age among tens of hundreds of possibilities for predictions, using cross-validation to compare learners with out-of-sample predictions. The ensemble SuperLearner can be used to combine baseline-knowledge from fixed epigenetic clocks like Horvath’s, while learning from new CpG sites with general machine learning algorithms. Using the discrete SuperLearner (the single best performing algorithm in the learner library) gives a standardized and conservative way to choose what epigenetic clock to implement for prediction. This is particularly appropriate for small sample sizes or if overfitting is a concern. Beyond DNA methylation, the SuperLearner framework provides a principled means of harmonizing aging metrics across diverse biological layers, including transcriptomic, proteomic, and telomeric data.

Our second contribution was reframing the evaluation of epigenetic aging in extraordinary regions like blue zones with a similar comparison population. Most studies rely on fixed clocks trained on many heterogenous cohorts, and are not designed to provide unbiased, well-tuned estimates in specific regions of interest. We trained a SuperLearner using only a representative sample of non-Nicoyan Costa Ricans. The ensemble relied mostly on the PCGrimAge and Hannum clocks, suggesting that existing clocks capture useful signal sufficient to predict age. We then used the comparison group’s age residuals to calibrate aging predictions in the Nicoyan blue zone population. Our calibration framework establishes an empirical way to test if a blue zone actually differs in biological age due to region, or if claims of healthier aging are the result of a poorly calibrated tool.

Among non-Nicoyan Costa Ricans, the SuperLearner stood out as the sole clock to indicate a marginal age deceleration or aging advantage of about a year (*R̄*_0_ = 1.09) (Figure 3). This implies that there may be aging advantages across Costa Rica. The uncalibrated analysis using existing epigenetic clocks illustrated the potential downfalls for the standard way epigenetic aging is evaluated. Depending on the chosen clock, Nicoyans had evidence to be anywhere from three years younger (SuperLearner, *R̄*_1_ = −3.05, 95% CI [−3.64, −2.46]) on average to almost four years biologically older (DNAmFitAge, *R̄*_1_ = 3.59, 95% CI [3.02, 4.17]) before calibration or adjustment. Once calibrating Nicoya to the reference population, all clocks converge to modest age deceleration by about one to two years, and SuperLearner still suggested the largest biological aging advantage (Δ̂ = 1.96, 95% CI [ 2.56, 1.37]). While all clocks had significant results, they suggest that living in Nicoya may not have the dramatic impacts on aging that we currently believe.

We adjusted Nicoyan aging predictions against confounders that impact the aging process: sex, body mass index, smoking and socioeconomic status (Appendix). In doing so, we saw that this adjustment had a very similar effect as calibration to Non-Nicoyan Costa Rica. Calibration to a well-matched population may help remove bias from unmeasured confounding in aging estimates. This is particularly important when confounders of interest cannot be observed, and also underscores that predicting epigenetic age is inherently difficult because the biological mechanisms of aging are themselves so intricate and not fully understood.

SuperLearner is designed to avoid overfitting with cross-validation, but this can still occur in finite samples. We also introduced a more rigorous way to align predictions with the comparison population using a subgroup (TMLE) calibration and mitigate overfitting by smoothing across groups. This strategy computes residuals in subgroups based on user-chosen variables to target that may shape epigenetic aging. We implemented the subgroup calibration using sex, a well-established effect modifier, and found little to no change in the results. In addition, we calibrated with true age as a subgroup to account for the stratified sampling process in the CRELES cohort. After splitting into 10-year age groups from 60 to 110, we found that most of the aging advantage vanished (Δ = 0.35, 95% CI [ 0.64, 0.05]). This is intuitive, especially in context of the true age vs predicted age plots for both populations with the SuperLearner (Figure 4). These plots suggest that most of the aging advantage across Costa Rica is captured in those about 90 years or older, and vanishes when you compare Nicoyans to Non-Nicoyans of similar age.

A visual summary of the results is best depicted in Figure 5, showing that the SuperLearner trained on non-Nicoyan Costa Ricans produced the most consistent estimates across estimation methods. Our framework can be extended to other longevity regions with data available on a matched comparison population to investigate and verify a more isolated effect of region on biological aging. In addition, this method can support other epigenetic aging questions involving estimating the burden, or benefit, of exposure. For instance, questions of whether exposure to environmental intoxicants effect epigenetic age can be investigated by using the average biological aging in a matched, representative reference sample as a calibration. It can be used similarly to estimate the impact of other well-established epigenetic aging contributors, like diet, exercise, or smoking. PCGrimAge contributed to the most ensemble SuperLearner, which targets physiological decline and is a strong predictor of mortality. Although longitudinal follow-up allows for mortality validation, the initial calibration remains sensitive to the fact that we cannot account for unobserved mortality and survivor bias. Future work should explore mortality as an outcome in addition to epigenetic age. In particular, whether these results remain stable when accounting for competing risks of death, or if the model overfits to the characteristics of the surviving population.

Research on the secrets of longevity in blue zones has been a hot topic for years. However, debates of blue zone validity have arisen, questioning whether individuals are actually living longer due to healthy aging, or if other factors are at play [30]. Some authors in non-peer reviewed press have raised concerns about age misreporting due to faulty or unknown birth and demographic records [31]. These concerns highlight the need for biologically grounded measures of aging that can be an additional way to validate blue zone longevity claims. Our empirical findings show a real but small biological aging advantage in Nicoya. Through targeted bias reduction with SuperLearner and calibration, we move toward inference to better understand the actual drivers of healthy aging.

## Data Availability

Public-use version of the CRELES data is available from the Inter-University Consortium for Political and Social Research (ICPSR) repository (https://www.icpsr.umich.edu/web/NACDA/studies/31263/versions/V1). Since data DNA methylation and the complementary sample of centenarians in Nicoya are not currently part of the public-use, requests for restricted access to data can be submitted at http://www.creles.berkeley.edu/ following institutional review approval.

## 5 Acknowledgements and Funding

This research was partially funded by the NIEHS Superfund Research Program (P42ES004705). Thank you to David Lin, Alexander Morin and Parmida Atashzay for technical contributions to DNA preparations.

## 6 Code/Data Availability

The software used to implement the DNAm SuperLearner is available at the following repository: https://github.com/nolangunter/DNAmSL. Public-use version of the CRELES data is available from the Inter-University Consortium for Political and Social Research (ICPSR) repository (https://www.icpsr.umich.edu/web/NACDA/studies/31263/versions/V1). Since DNA methylation data and the complementary sample of centenarians in Nicoya are not currently part of the public-use, requests for restricted access to data can be submitted at http://www.creles.berkeley.edu/ following institutional review approval.

## 7 Appendix

### 7.1 Epigenetic aging residuals adjusted for important confounders

Epigenetic aging estimates are commonly not just derived from raw residuals of predictions or running a simple linear regression with the predictions, but regressed against an additional confounder set to remove any explainable bias. We use a confounder set of biological sex, body mass index (BMI), and continuous measures of smoking status and socioeconomic status. For the fixed epigenetic clocks, we may use the confounder set in a linear regression as normal. However, to avoid double regression and variance blowup with the SuperLearner model, we explore two alternatives. We propose two ways to adjust for the confounder set with SuperLearner predictions:

- **Residual (R)-Adjusted SL**: Run an initial regression of age against the confounder set, then use the residuals as the outcome to train a new SuperLearner.
- **Direct (D)-Adjusted SL**: We trained an adjusted SuperLearner by including the four confounders as predictors in the candidate sets (alongside CpGs) picked by SuperLearner. Thus, the variables wouldn’t appear for any fixed epigenetic clocks, but could appear in the flexible machine learning algorithms.

Table 4 shows the results for running an Ensemble SuperLearner on each version. The Direct-Adjusted SL still puts most of it’s weight into the PCGrimAge clock (73.7%), but actually does put more weight into flexible machine learning algorithms with more weight in ridge regression. This suggests that it picked up more information from including the confounders in the predictor set. The Residual-Adjusted SL has a dramatic shift from the ensemble weights we’ve seen before, now putting the majority of weight in flexible machine learning algorithms. Big Lasso was given 72.2% weight alongside other ridge regression and gradient boosting learners. Using the confounder-adjusted residuals as the outcome allowed the SuperLearner to pick up on new information in the CpG predictor set.

Finally, we show the results of the confounder-adjustment for each clock in Table 5. The point estimates of epigenetic aging in Nicoya and associated confidence intervals are close to the calibrated results in Table 3. Still, the SuperLearner estimates the highest epigenetic aging advantage, between about two to two-and-a-half epigenetic years younger. The standard deviation of the Residual-Adjusted learner is inflated due to double regression, but the effect is still significant, (*R̄*_1_ = 2.10, 95% CI [ 2.91, 1.29]). The Direct-Adjusted SuperLearner maintains a stable standard deviation and also finds a significant effect (*R̄*_1_ = 2.59, 95% CI [ 3.19, 2.00]). The comparison fixed epigenetic clocks also are all significant, but on average estimate about one to two years of an aging advantage among Nicoyans.

### 5.2 Subgroup (TMLE) Calibration Results

While calibration by the mean residuals in the comparison population may be sufficient, it is also important to account for additional sources of variation that influence DNA-methylation–based age estimates. We implement a targeted maximum likelihood–style calibration to correct systematic bias from effect modifiers in Super Learner predictions, extending the work from Brooks et al [32]. It is a form of calibration based on subgroups of interest, aiming to increase robustness by smoothing and mitigating any overfitting in the model. Let *f̂*_0_(*W* ) denote the initial cross-validated predictions fit on the reference population, and let *Y* denote the true chronological age. We implement a procedure to reduce residual bias that differs across a variable (or variables) Z. For example, we may partition the continuous age range into discrete categories = *z*_1_*, . . . , z*_K_ using pre-specified age breaks, assigning each individual to an age bin *Z*. This naturally follows with binary or categorical variables, like biological sex. Given these categories, we fit a fluctuation model with an offset equal to the initial predictions:

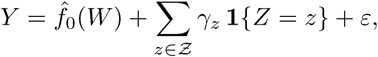

where *γ*_z_ represents a stratum-specific correction term estimated by ordinary least squares. The updated, targeted prediction is then:

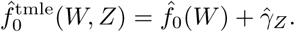

This simple fluctuation step updates the initial model predictions to match the observed age means within each stratum while preserving the overall structure learned by the Super Learner. By construction, the targeted update brings 𝔼̂ [*f̂*_0_(*W* ) *Z* = *z*] closer to 𝔼̂ [*Y Z* = *z*] for each stratum, yielding a stratified calibration that aligns predicted and observed values. This procedure corresponds to the canonical TMLE fluctuation step under an identity link with offset *f̂*_0_(*W* ). In practice, the standard implementation is to fit a generalized linear model in which chronological age is regressed on the stratum, using the initial predictions from Super Learner as an offset in the model. It represents a minimal one-step correction that can later be extended to continuous calibration functions, higher-order targeting, or logistic links for bounded outcomes. Typically, men have higher epigenetic age acceleration and weaker correlations between chronological and methylation-predicted age than women [33]. However, in most blue zones, particularly in Sardinia and Nicoya, men show exceptional longevity compared to women [6, 15], making biological sex a significant effect modifier. To test for sex-specific calibration bias, we implement a subgroup-calibrated targeted update using biological sex as the sole covariate. This allows the estimator to flexibly adjust for systematic residual differences, thereby aligning the calibrated methylation-age scale across men and women. We rerun the one-sample t-test for each clock with residual adjustments stratified by sex (Table 6). Incorporating biological sex into the TMLE calibration yielded nearly identical calibrated predictions to the mean-residual calibration, indicating that sex contributed minimal additional bias correction within the comparison group of Non-Nicoyans. Thus, the sex-specific offset term did not materially alter the calibrated methylation-age estimates.

We also run a similar calibration adjusting for an age-specific offset term instead of sex. Since Nicoyans over 95 years were oversampled, we want to account for a difference in epigenetic aging that may come from selection bias. In particular, only individuals living long enough to be sampled at a higher age are observed. Naturally, these individuals likely have an epigenetic aging advantage compared to their deceased peers, who are not measured. We compute an overall difference in epigenetic age by calibrating to the differences in 10-year age bins. The residual age difference estimates become much smaller in magnitude with this adjustment. While most of the fixed epigenetic clock estimates are no longer significant, PCGrimAge(Δ̂ = −0.50, 95% CI [−0.94, −0.06]) and SuperLearner (Δ̂ = −0.35, 95% CI [−0.64, −0.05]) still suggest a significant but small aging advantage among Nicoyans.

### 5.3 Impact of Calibration on Variance and Power

If the clock were trained on pooled data without calibration (as is typical for fixed clocks), the expected residual in the reference population need not be zero, i.e., E[*R*_0_] = 0. A naive comparison would therefore require a two-sample *t*-test between the mean residuals of both groups:

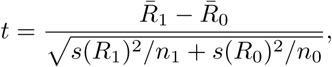

which tests for differences after absorbing whatever baseline bias the global model introduces. To illustrate how calibration improves this comparison, consider a simple linear data-generating process within each population *A* ∈ {0, 1}:

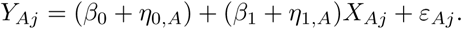

Here:

- *Y*_Aj_ is chronological age for person *j* in population *A*,
- *X*_Aj_ is a summary of DNA methylation (or a linear score based on *W*_Aj_),
- *η*_0,A_ and *η*_1,A_ represent population-specific shifts in intercept and slope,
- *ε*_ij_ is individual noise with variance *σ*^2^.

A global clock *f*_pool_ trained on pooled data ignores the group-specific terms (*η*_0A_*, η*_1A_). The resulting prediction error includes both individual noise and population heterogeneity:

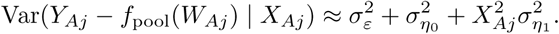

Thus, comparing residuals from a pooled model confounds biological aging differences with between-population regression mismatches. By contrast, a reference-calibrated clock, trained only on the reference group (*A* = 0) and mean-centered to satisfy E[*R*_0_] = 0, absorbs the intercept shift *η*_0,0_. Its residual variance in the reference population reduces to:

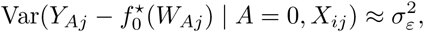

i.e., the variance attributable purely to individual-level aging noise. Because calibration eliminates between-population error in the reference group, the variance of the simple estimator Δ̂ = *R̄*_1_ is smaller than the variance arising from the pooled two-sample comparison. Lower variance yields greater power to detect a true population-level deviation Δ *<* 0 (blue-zone advantage). Thus, calibration removes nuisance variation due to population differences in methylation–age structure, ensuring that any residual shift in the blue-zone population reflects a genuine deviation in biological aging rather than a modeling artifact.

## Footnotes

1 https://populationsciences.berkeley.edu/creles/

